# Hemodynamic Instability and Retinal Vein Occlusion in Glaucoma: Comparative Analysis of Heart Rate Variability and Choroidal Perfusion

**DOI:** 10.1101/2025.04.23.25326269

**Authors:** Ji Hye Lee, Young-Hoon Park

**Author notes:** **Address for Correspondence:** Young-Hoon Park, MD, PhD, Department of Ophthalmology and Visual Science, Seoul St. Mary’s Hospital, College of Medicine, The Catholic University of Korea, Seoul, Republic of Korea, 222 Banpo-daero, Seocho-gu, Seoul 06591, Republic of Korea. **Financial Disclosure:** none. **Conflict of Interest:** none.

## Abstract

**Purpose:** To assess the hemodynamic and structural differences between glaucoma patients who developed retinal vein occlusion (RVO) and those who did not.

**Study Design:** Retrospective, single-center, cross-sectional study

**Methods:** This study included glaucoma patients who underwent a heart rate variability (HRV) test between January 2018 and July 2024. Patients were subdivided into RVO and non-RVO groups. Baseline mean deviation (MD) and pattern deviation (PSD) of the visual field and optical coherence tomography parameters were analyzed.

**Results:** Twenty-nine glaucoma patients with RVO and 34 glaucoma patients without RVO were included. Baseline MD and PSD had no difference (MD: −5.98 ± 9.05 vs. −3.70 ± 4.70, *p*=0.114; PSD: 3.88 ± 3.78 vs. 3.98 ± 3.96, *p*=0.883). HRV parameters, specifically Standard Deviation of NN interval (SDNN) and root-mean-square of successive differences (rMSSD), were significantly lower in the RVO group (SDNN: 22.12 ± 8.27 vs. 36.71 ± 24.74, *p*=0.002; rMSSD: 16.34±9.55 vs. 29.87±31.58, *p*=0.022). Statistically significant choroidal vascularity index difference was also observed between groups (64.62±7.38 vs. 67.49±5.90, *p*=0.045). Multivariate logistic regression analysis identified hypertension medication use (*p*=0.019) and SDNN (*p*=0.003) as factors associated with RVO development in glaucoma patients.

**Conclusion:** This is the first reported study to analyze the association between hemodynamic instability and RVO development in glaucoma patients. Our results showed that RVO patients had lower HRV parameters, with SDNN being significantly associated with RVO development. We believe that HRV can be an indicator for identifying glaucoma patients at risk for RVO, though further studies are needed to validate this association.

## Introduction

Despite the development of many treatment options, such as intravitreal anti-vascular endothelial growth factor (anti-VEGF) agents, steroid injection, and laser treatment, retinal vein occlusion (RVO) remains the second most common retinal vascular disease after diabetic retinopathy, and is a leading cause of severe vision loss (1, 2). RVO can be classified based on the site of occlusion: branch RVO occurs mainly at an arteriovenous intersection, and central RVO occurs at or near the lamina cribrosa of the optic disc (1, 3). RVO is associated with various systemic risk factors, including advanced age, hypertension, diabetes mellitus, dyslipidemia, atherosclerosis, and blood viscosity (4–7). In addition to these systemic risk factors, glaucoma is the primary ocular risk factor for RVO (6, 8–10). Shin et al.(11) reported that autonomic dysfunction can cause blood flow abnormalities in glaucoma patients, potentially accelerating disease progression. This hemodynamic instability could contribute to both the progression of glaucoma and the development of RVO, as RVO is closely related to systemic vascular health.

This study aims to identify the risk factors in glaucoma patients who are more likely to develop RVO and investigate whether autonomic dysfunction in glaucoma patients could increase the risk of RVO development.

## Methods

### Ethics statement

This retrospective, cross-sectional study was approved by the Institutional Review Board (IRB) of Seoul St. Mary’s Hospital, Catholic University of Korea (KC24RASI0497), and adhered to the tenets of the Declaration of Helsinki. As the study was conducted retrospectively, the local IRB of the Catholic University of Korea waived the need for informed consent.

### Participants

We retrospectively reviewed the medical records of glaucoma patients who underwent heart rate variability (HRV) testing between January 2018 and July 2024. Patients were subdivided into two groups: those who developed RVO during the follow-up period and those who did not. Patients with concurrent ocular diseases that could affect the functional and anatomical status, such as optic nerve disease, advanced age-related macular degeneration (AMD), diabetic retinopathy, high myopia with axial length ≥ 26 mm, and poor image quality, were also excluded.

All patients underwent complete ophthalmic evaluations, including best-corrected visual acuity, Goldmann applanation tonometry, fundoscopy, swept-source optical coherence tomography (SS-OCT) (DRI Triton, Topcon, Tokyo, Japan), and Humphrey visual field (VF) examination using the Swedish interactive threshold Standard 24-2 algorithm (Carl Zeiss Meditec, Dublin, CA, USA). Patients were also referred to the internal medicine department for HRV tests at the time of glaucoma diagnosis, and their medical histories were thoroughly reviewed.

### Measurements

The 3 mm diameter inner ring and 6 mm diameter outer ring were segmented into superior, inferior, nasal, and temporal quadrants, according to the Early Treatment of Diabetic Retinopathy Study (ETDRS) macular map sectors (12). Central macular thickness (CMT), ganglion cell-inner plexiform layer (GGIPL) thickness, and each sectoral retinal thickness were measured using the OCT software.

Choroidal thickness (CT) was measured using the built-in calipers of the OCT device. Subfoveal CT was determined as the distance between the outer border of the retinal pigment epithelium (RPE) and the inner border of the suprachoroidal space (13). To minimize inter-observer variability, two retinal specialists (J.H.L. and Y.H.P.) measured CT independently, and the average value was used.

The choroidal vascularity index (CVI) was evaluated to assess the vascularity status of the choroid. The selected image was segmented following the protocol described by Agrawal et al., and image binarization was performed using the ImageJ 1.54 software (National Institute of Health, Bethesda, MD, USA)(14, 15). Using the polygonal selection tool, the subfoveal choroidal area with a width of 1,500 microns, demonstrating the total subfoveal circumscribed choroidal area (TCA), was selected and then added to the regions of interest (ROI) manager. The image was converted into 8-bit, and a Niblack auto local threshold was applied. After applying the color threshold tool, the stromal area (SA) was added to the ROI manager. The TCA and SA were merged using an “AND” operation within the ROI manager, creating a third area. The luminal area (LA) was calculated as the difference between the TCA and the third area. The CVI was calculated as the ratio of LA to TCA (CVI = LA/TCA × 100) (Figure 1).

**Fig 1.**
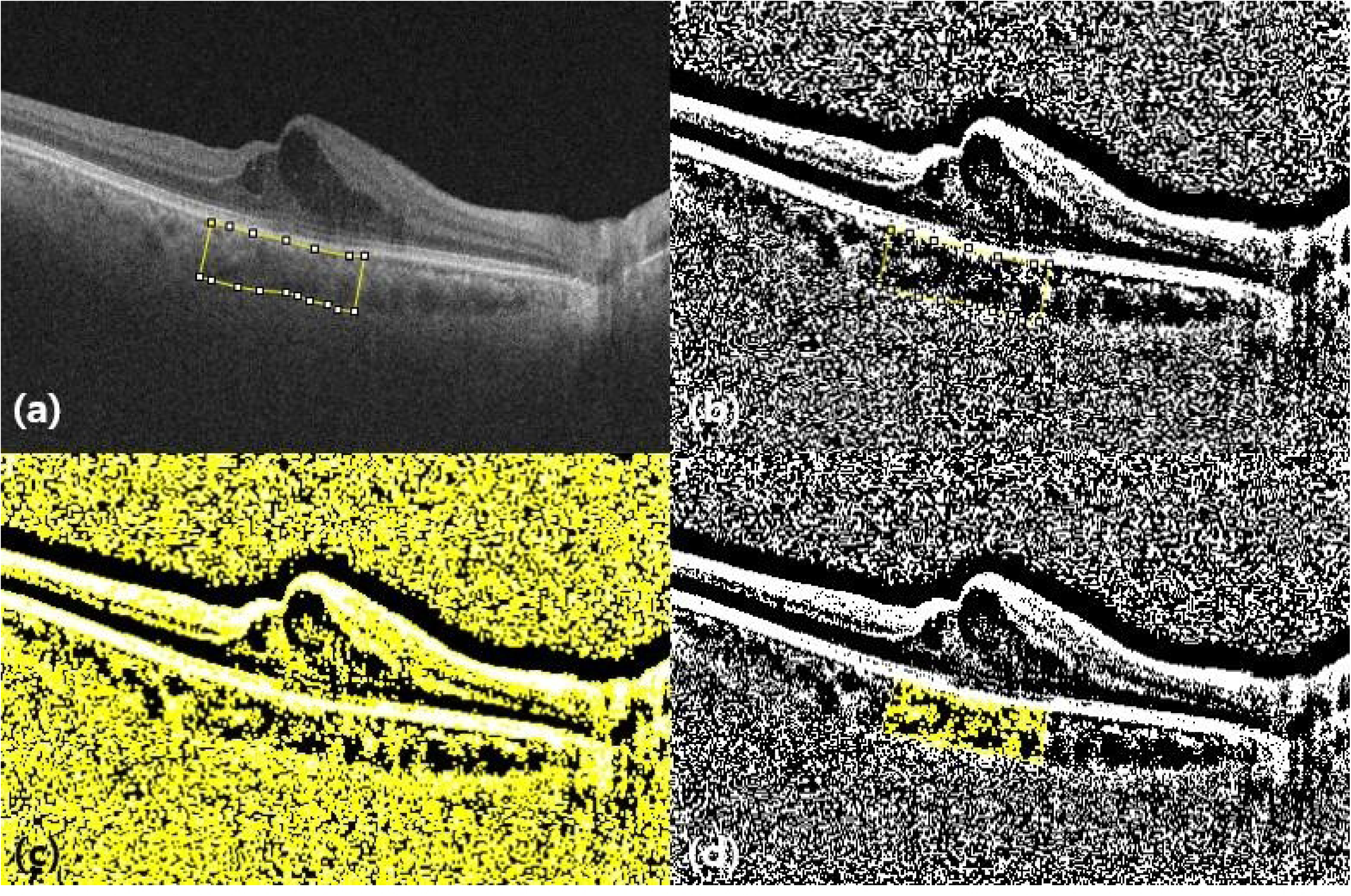
Choroidal vascularity index (CVI) measurement using Image J software. **a.** OCT image with 1,500 µm segmentation block of the subfoveal choroidal area. **b.** Image binarization using the Niblack autolocal threshold. **c.** Applying the color threshold tool, added the stromal area to the region of interest (ROI) manager. **d.** CVI was calculated as the ratio of luminal area (LA) to total choroidal area (TCA).

### HRV assessment

Two time-domain HRV indices were retrieved: the standard deviation of all normal-to-normal R-Rs intervals (SDNN), representing the total effects of the regulation of the autonomic nervous system, and the root mean square of successive differences between normal-to-normal R-R intervals (rMSSD), which measures short-term variation and thus estimates the high-frequency variations in heart rate and is thought to be highly correlated (16–18). Reduced SDNN values indicated an autonomic imbalance. The average SDNN and rMSSD values for Koreans in their 60s were reported as approximately 38 ms and 23 ms, respectively (16, 19).

The frequency-domain analysis included the absolute and relative powers of the low frequency (LF, 0.04–0.15 Hz), high frequency (HF, 0.15–0.4 Hz), and LF/HF ratio. The LF and HF measurements represent the sympathetic and parasympathetic modulations, respectively; thus, the LF/HF ratio depicts the overall sympathovagal balance (19). The median LF/HF ratio among Koreans in their 60s–70s was 2.91–3.61 (19).

### Statistical Analyses

Statistical analyses were performed using the SPSS software (version 24.0; IBM Corp., Armonk, NY, USA). An independent t-test was performed to compare the quantitative measures between glaucoma patients with and without RVO, while Pearson’s chi-square test was used to compare the baseline demographics of the patients. A paired t-test was performed to compare the inter-eye differences in RVO patients. Logistic regression analysis was performed to assess the predictive factors associated with the development of RVO in glaucoma patients. Variables with a significance level of *p* <0.20 in the univariate analysis were included in the multivariate model. Statistical significance was set at *p* <0.05.

## Results

A total of 63 patients diagnosed with glaucoma were included in this study, of whom 34 developed RVO. Table 1 presents the baseline demographic and ophthalmic characteristics of patients. No statistically significant differences were observed between the two groups in baseline intraocular pressure and VF indices, including MD and PSD. However, HRV parameters showed differed significantly, with patients who developed RVO showing lower SDNN and rMSSD values compared to those without RVO (SDNN: 22.12 ± 8.27 vs. 36.71 ± 24.74, *p* = 0.002; rMSSD: 16.34 ± 9.55 vs. 29.87 ± 31.58, *p* = 0.022) (Table 1).

**Table 1.** Baseline Characteristics and Comparisons of Glaucoma Patients According to the Development of RVO.

p values were calculated using independent t-test (\*) and Pearson's Chi-square test (†)
|  | Glaucoma with RVO<br>(n=29) | Glaucoma without RVO<br>(n=34) | P value |
| --- | --- | --- | --- |
| Age (years) | 66.03 ± 9.86 | 64.79 ± 10.70 | 0.636 † |
| Sex, male:female, n | 13:16 | 13:21 | 0.596 † |
| Medication of DM, n (%) | 9 (31.0 %) | 5 (14.7 %) | 0.120 † |
| Medication of HTN, n (%) | 17 (58.6 %) | 14 (41.2 %) | 0.167 † |
| Baseline IOP (mmHg) | 14.78 ± 3.73 | 15.28 ± 4.24 | 0.484 † |
| Baseline MD of VF (dB) | -5.98 ± 9.05 | -3.70 ± 4.70 | 0.114 * |
| Baseline PSD of VF (dB) | 3.88 ± 3.78 | 3.98 ± 3.96 | 0.883 * |
| HR (bpm) | 70.07 ± 14.17 | 67.91 ± 10.47 | 0.491 * |
| SDNN (ms) | 22.12 ± 8.27 | 36.71 ± 24.74 | **0.002** * |
| rMSSD (ms) | 16.34 ± 9.55 | 29.87 ± 31.58 | **0.022** * |
| LF/HF ratio | 1.40 ± 1.39 | 1.59 ± 1.53 | 0.599 * |
| Follow-up period (years) | 8.00 ± 5.11 | 9.00 ± 5.20 | 0.446 † |

Statistically significant differences between two groups ( $P < 0.05$ ) are indicated in bold

### Optical coherence tomography parameters

We compared OCT parameters between patients with RVO and those without vascular changes. Retinal and GCIPL thicknesses measured at the acute stage of RVO in affected eyes showed significant differences between the two groups (Table 2). RVO patients had significantly lower CVI compared to those without RVO (64.62 ± 7.38 vs. 67.49 ± 5.90, *p* = 0.045). However, no significant difference was observed in subfoveal CT (241.24 ± 81.15 vs. 258.68 ± 84.71, *p* = 0.350) (Table 2). Additionally, when comparing the structural components between the fellow eye of the RVO patients, which is the non-affected side, and non-RVO patients, no statistically significant differences were found (Table 2-1).

**Fig 2.**
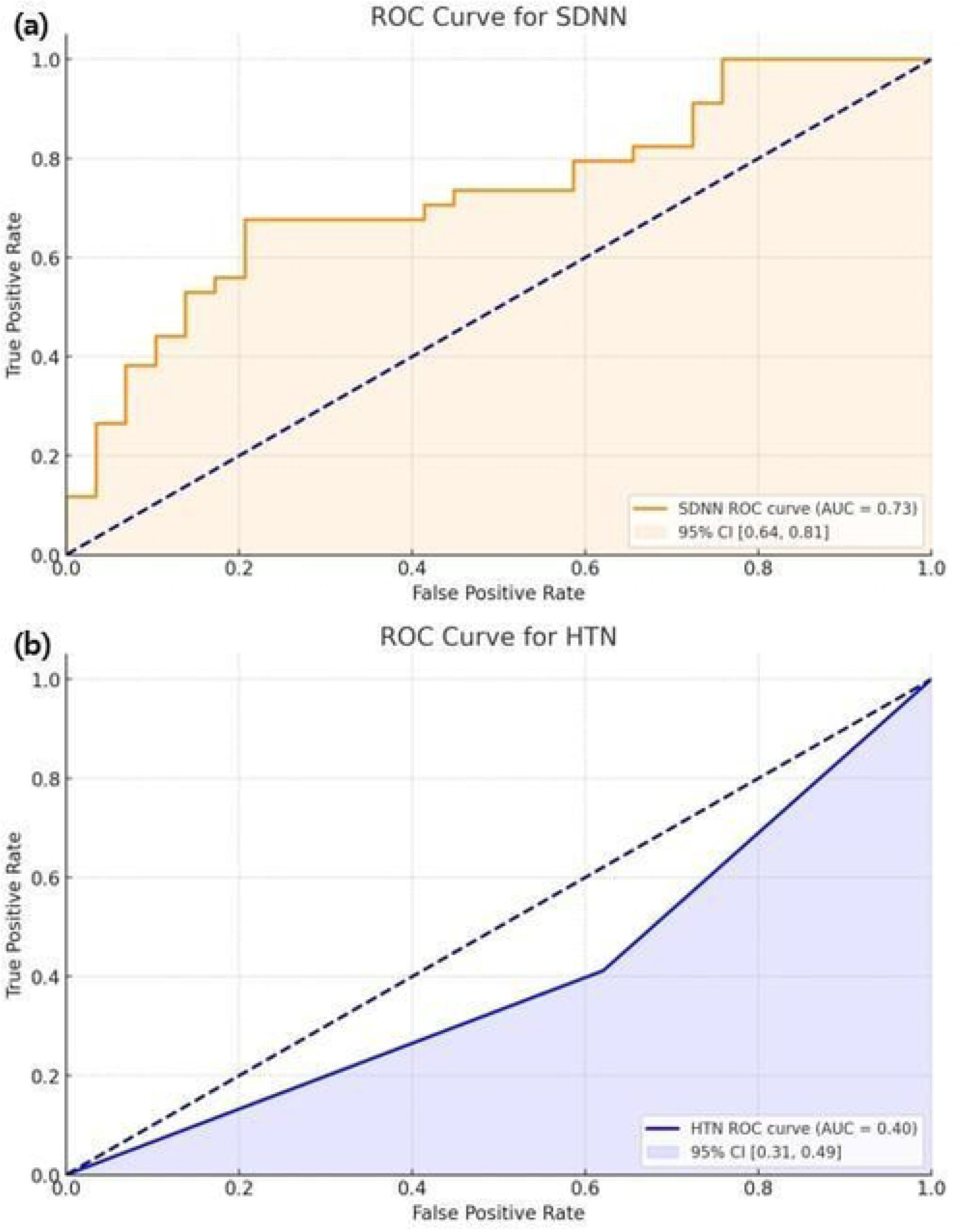
Receiver operating characteristic curve (ROC) for the prediction of retinal vein occlusion (RVO) based on the standard deviation of NN interval (SDNN) and hypertension (HTN).

**Table 2.**
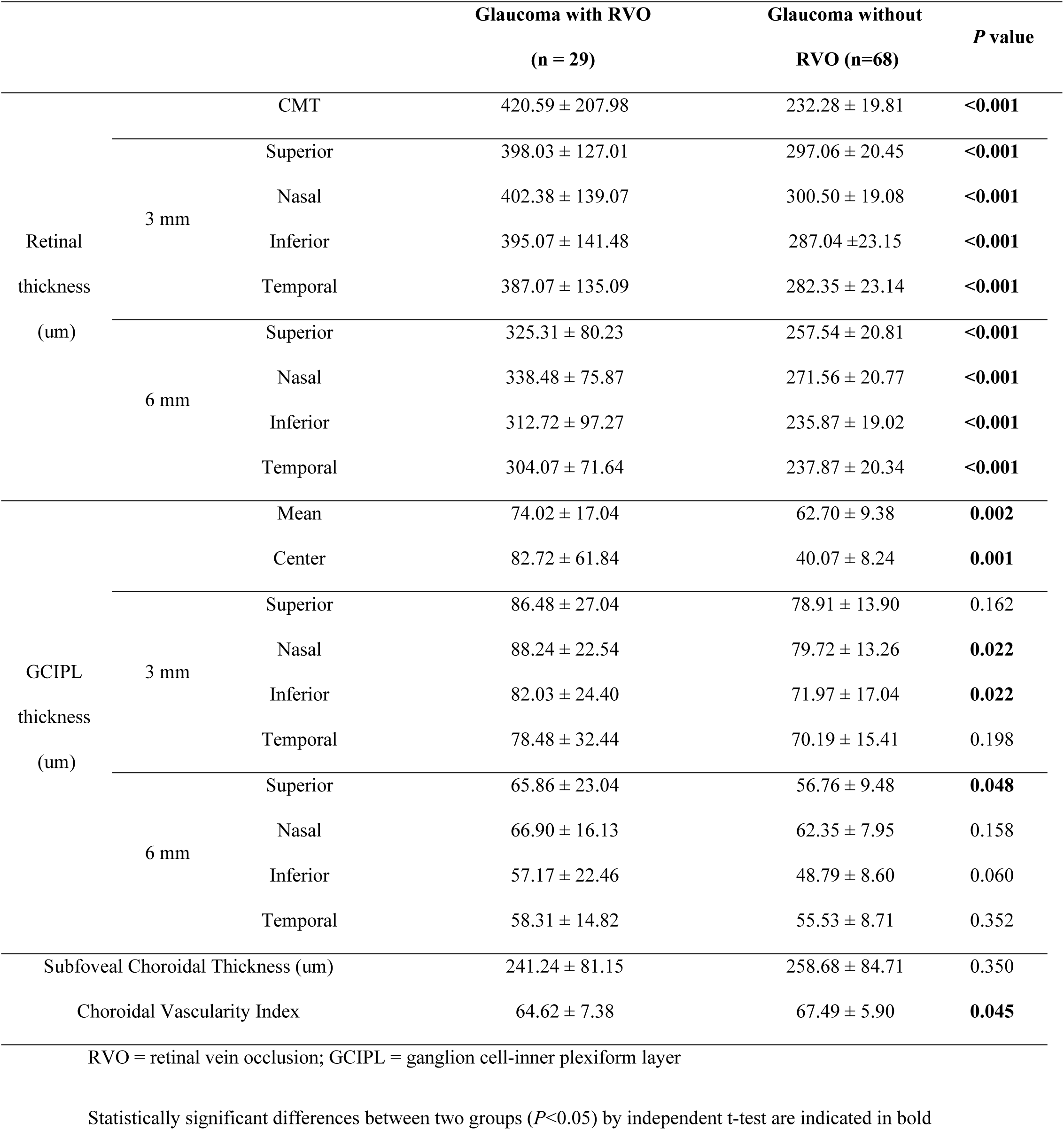
Comparison between OCT parameters in the affected eyes of patients with Retinal vein occlusion and controls.

**Table 2-1.**
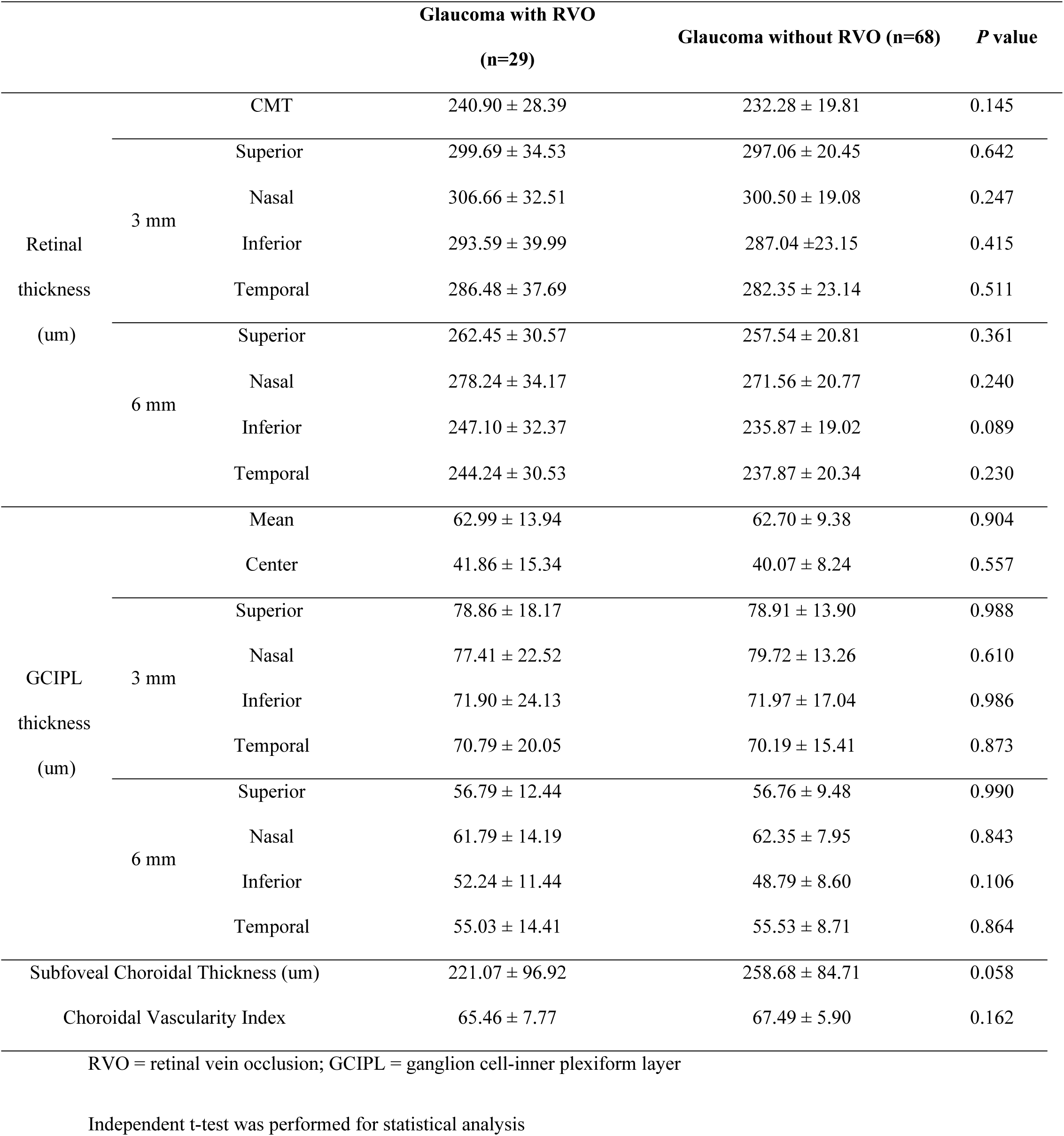
Comparison between OCT parameters in the fellow eyes of patients with retinal vein occlusion and controls.

### Inter-eye measurements

In the group analysis of RVO patients, the affected and unaffected eyes showed no functional differences in the VF test, but significant differences were observed in retinal thickness (Table 3). Mean GGIPL thickness, as well as center and nasal GCIPL thickness within the 3 mm diameter inner ETDRS grid, differed significantly between the affected and unaffected eyes (*p* = 0.005, *p* = 0.002, *and p* = 0.045, respectively). The inter-eye comparison of subfoveal CT and CVI showed no statistically significant differences (CT: 221.07 ± 96.92 vs. 241.24 ± 81.15, *p* = 0.574; CVI; 65.46 ± 7.77 vs. 64.62 ± 7.38, *p* = 0.196).

**Table 3.**
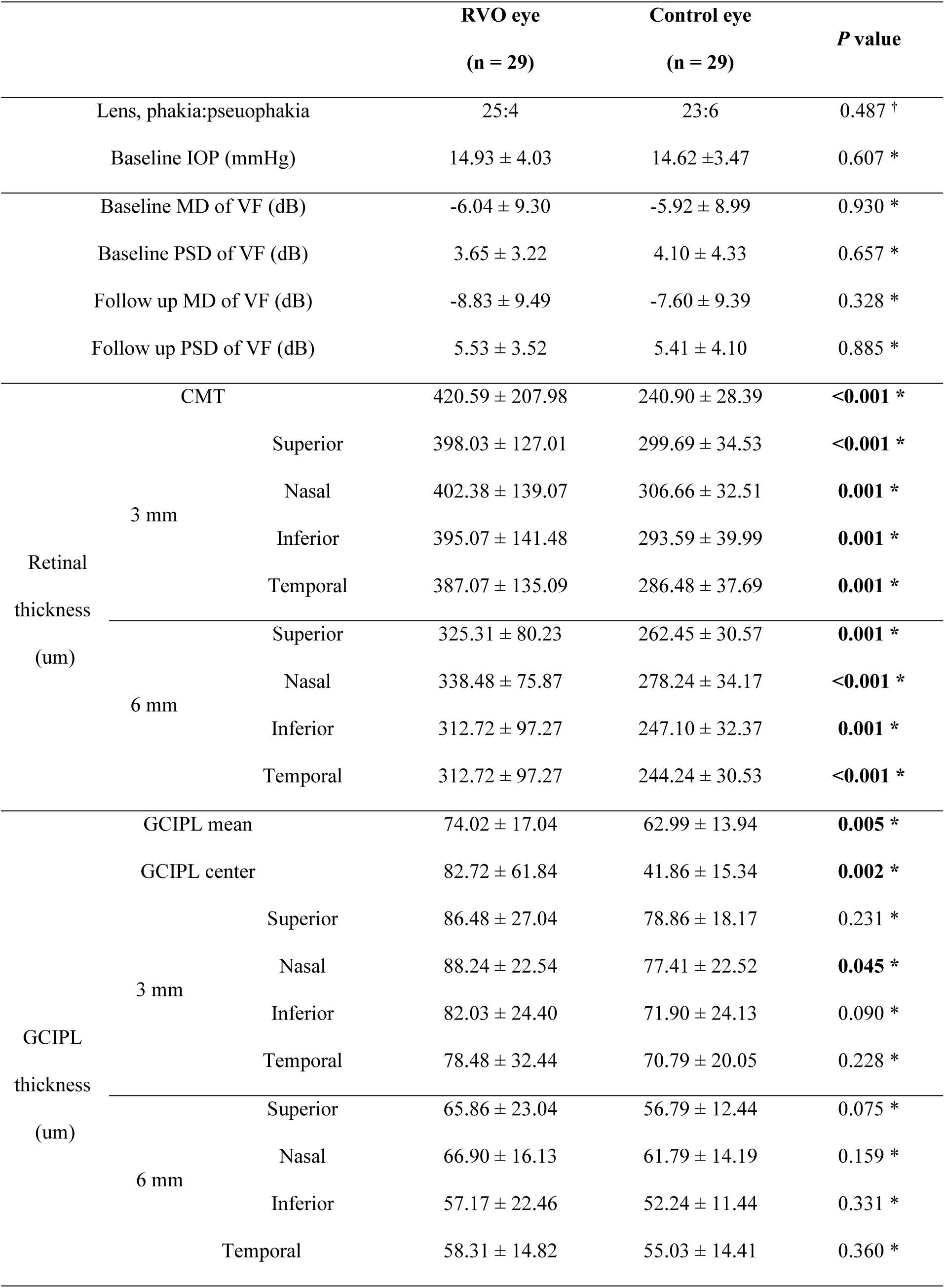
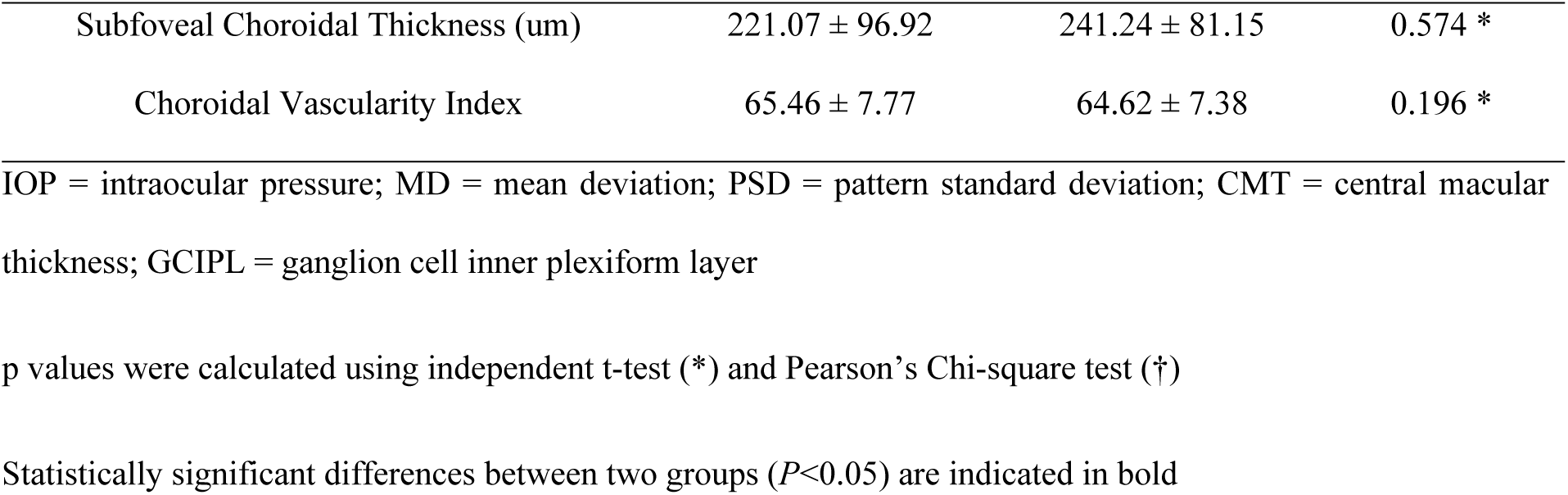
Inter-eye difference in patients with retinal vein occlusion.

### Predictive factors affecting the development of RVO

Factors associated with the development of RVO in glaucoma patients were analyzed (Table 4). Medication for diabetes (*p* = 0.120), systemic hypertension (*p* = 0.098), baseline MD (*p* = 0.085), subfoveal CT (*p* = 0.081), CVI (*p* = 0.049), and HRV parameters, including SDNN (*p* = 0.008) and rMSSD (*p* = 0.057) were significantly associated with RVO development in the univariate analysis. Among these factors, SDNN emerged as the predictive HRV factor for developing RVO in glaucoma patients (odds ratio [OR], 1.137; 95% confidence interval [CI], 1.044–1.238; *p* = 0.003). Systemic hypertension was also identified as a significant risk factor for RVO development (OR, 0.321; 95% CI, 0.124–0.831, *p* = 0.019). Receiver operating characteristic (ROC) curve analysis demonstrated that the SDNN has a moderate predictive power, with an area under the ROC curve (AUC) of 0.73 (95% CI: 0.64–0.81), while systemic hypertension had relatively low predictive power for RVO occurrence (AUC: 0.40, 95% CI: 0.31–0.49). (Figure 2).

**Table 4.** Factors Associated with the Development of retinal Vein Occlusion.

MD = mean deviation; PSD = pattern standard deviation; VF = visual field; CT = choroidal thickness; CVI =
| Variables | Univariate |  | Multivariate |  |
| --- | --- | --- | --- | --- |
|  | OR (95% CI) | *P* value | OR (95% CI) | *P* value |
| Age (years) | 0.988 (0.941-1.038) | 0.630<sup>†</sup> |  |  |
| Sex, male:female, n | 1.313 (0.479–3.593) | 0.596* |  |  |
| Medication of DM | 0.383 (0.112-1.314) | **0.120\*** | 1.097 (0.361-3.337) | 0.871 |
| Medication of HTN | 0.428 (0.155-1.180) | **0.098\*** | 0.321 (0.124-0.831) | **0.019** |
| Baseline MD of VF (dB) | 1.051 (0.993-1.112) | **0.085<sup>†</sup>** | 1.061 (0.991-1.137) | 0.088 |
| Baseline PSD of VF (dB) | 1.008 (0.915-1.109) | 0.875<sup>†</sup> |  |  |
| Subfoveal CT (um) | 1.004 (1.000-1.008) | **0.081<sup>†</sup>** | 1.001 (0.996-1.006) | 0.632 |
| CVI | 291.249<br>(1.018-83329.762) | **0.049<sup>†</sup>** | 136.603<br>(0.225-82900.951) | 0.133 |
| HR (bpm) | 0.985 (0.946-1.027) | 0.485<sup>†</sup> |  |  |
| SDNN (ms) | 1.094 (1.023-1.170) | **0.008<sup>†</sup>** | 1.137 (1.044-1.238) | **0.003** |
| rMSSD (ms) | 1.055 (0.998-1.114) | **0.057<sup>†</sup>** | 0.943 (0.886-1.004) | 0.065 |
| LF/HF ratio | 1.100 (0.776-1.560) | 0.593<sup>†</sup> |  |  |
| Follow-up period (years) | 1.039 (0.942-1.146) | 0.439<sup>†</sup> |  |  |

choroidal vascularity index; HR = heart rate; SDNN = standard deviations of NN interval; rMSSD = root-mean-square of successive differences; LF = low frequency; HF = high frequency
p values were calculated using Pearson's Chi-square test (\*) and logistic regression analysis (†)
Factors with $P < .2$ in univariate analysis were included in multivariate analysis

## Discussion

Previous studies have demonstrated the relationship between glaucoma and RVO development. In glaucoma patients, RVO is believed to arise from glaucomatous structural changes, such as modifications at the lamina cribrosa, or it may coexist with retinal hemodynamic instability (9, 20). While glaucoma alone can cause severe visual dysfunction, the onset of RVO can accelerate this deterioration, resulting in irreversible ophthalmic impairment. Thus, identifying risk factors and focusing on preventing RVO in glaucoma patients is critical.

Our study demonstrated that glaucoma patients taking systemic hypertension medications were more likely to develop RVO. Additionally, patients with low SDNN, which represents dysregulation of the autonomic nervous system, showed a higher tendency to develop retinal vein occlusion. Cardiovascular assessment of autonomic dysfunction via HRV is widely used because it is non-invasive and relatively easy to implement (21). Previous research has shown that patients with low HRV had a higher risk of developing diabetic retinopathy (17). Ocular blood flow is regulated by both direct and indirect mechanisms of the autonomic system (22). Two separate vascular systems, retinal and uveal, primarily represented as choroidal vasculature, are influenced by autoregulatory influences, either indirectly or directly (22). However, the precise impact of the autonomic nervous system on retinal vasculature remains ambiguous, highlighting the importance of systemic hemodynamic changes (23). Autonomic dysfunction disrupts the balance of sympathetic and parasympathetic tone, leading to systemic imbalances in vasoconstriction and vasodilatation (23, 24). Our results demonstrated that patients who developed RVO had lower SDNN and rMSSD at baseline. This suggests that RVO patients may have a vulnerable autonomic nervous system, leading to an imbalance in vascular constriction and dilation. This dysfunction, similar to that seen in diabetic retinopathy, may ultimately contribute to the development of retinopathy.

Several retinal vascular diseases, including diabetic retinopathy, age-related macular degeneration, and retinopathy of prematurity, involve choroidal vascular changes that either accompany or precede retinal vascular alterations (25, 26). However, choroidal involvement in RVO remains unclear, with conflicting evidence on whether eyes affected by RVO have higher or lower CVI compared to their fellow eyes (14, 25, 27). Some studies suggest that impaired retinal venous outflow in RVO could cause extracellular fluid to shift towards the choroid, leading to subsequent choroidal stromal swelling and congestion. Aribas et al.(25) reported that compared with healthy control eyes, the non-affected fellow eyes of the RVO patients had lower CVI. Our results showed no statistically significant differences in inter-eye comparison of CT and CVI values between the affected and fellow eyes of RVO patients (Table 3). However, when comparing the CVI of RVO-affected eyes to that of the control participants, who have glaucomatous eyes without RVO, the RVO-affected eyes had significantly lower CVI (Table 2). Additionally, the fellow eyes of RVO patients tended to have lower CVI compared to the control participants (Table 2-1). RVO is a condition strongly influenced by systemic vascular health. The decreased CVI observed in RVO patients compared to non-RVO patients in our study indicates alterations in choroidal vessels, which are partially associated with systemic vascular disease. Since glaucoma patients typically exhibit reduced CVI compared to healthy eyes, may have contributed to the lack of a statistical difference in CVI between RVO-affected eyes and their non-affected fellow eyes (28).

This study had several limitations. First, due to the limited number of glaucoma patients who underwent the HRV test and eventually developed RVO, we could not perform a subgroup analysis based on the type of glaucoma, such as normal-tension or primary open-angle glaucoma. Further studies with larger samples are warranted to obtain higher statistical power.

Another limitation is that HRV results can be affected by antihypertensive drugs such as angiotensin-converting enzyme inhibitors (ACE) and ß-blockers, which could increase high-frequency HRV and reduce low- and mid-frequency bands, respectively (29). While we investigated the patients’ underlying metabolic diseases, we could not determine the specific medications that they were currently taking. This underlying drug history may have affected the HRV results.

### Conclusion

HRV is a non-invasive and relatively easily accessible method of detecting autonomic dysfunction.

To our knowledge, this is the first reported study to investigate whether autonomic dysfunction, which is represented by low HRV test values, is a risk factor for the development of RVO in glaucoma patients. Although the exact nature of this relationship needs further clarification, glaucoma patients with low HRV should be closely monitored for retinal vascular diseases. We believe that HRV can be an indicator of susceptibility to RVO in glaucoma patients, though further studies are needed to clarify this association.

## Data Availability

All relevant data are within the manuscript and its Supporting Information files.

## Acknowledgments

This study was supported by the Basic Science Research Program through the National Research Foundation of Korea (NRF-RS-2023-00253065). The funders had no role in study design, data collection and analysis, decision to publish, or preparation of the manuscript.

